# Individual-Level Double Burden of Malnutrition among Ethiopian Women: Prevalence, Machine-Learning Prediction and Explainability Using the 2024/25 Ethiopian Demographic and Health Survey

**DOI:** 10.64898/2026.09.22.26363745

**Authors:** Ashebir Mamay Gebiru, Geta Bayu Genet, Solomon Tibebu Ambelie, Serku Abate Mihret, Demeke Wondie Azene, Mulualem Endeshaw Zeleke, Aklilu Yiheyis Abereha, Yadelew Yimer Shibabaw

## Abstract

**Background:** The double burden of malnutrition, combining overweight or obesity with anemia is vital in nutritional transitions. Evidence regarding its national distribution and prediction using routine survey data in Ethiopia remains limited.

**Objective:** To estimate the national prevalence of individual-level double burden of malnutrition among non-pregnant Ethiopian women aged 15-49 years and evaluate machine-learning models using data from the 2024/25 Ethiopian Demographic and Health Survey.

**Methods:** We analyzed 14250 non-pregnant women. Double burden of malnutrition was defined as a body mass index equal to or greater than 25.0 kilograms per square meter with an altitude- and smoking-adjusted hemoglobin concentration below 12.0 grams per deciliter. Data were split into an 80% training set and a 20% holdout test set. Six algorithms were evaluated using cross-validation, while SHapley Additive exPlanations described feature contributions.

**Results:** The weighted national double burden of malnutrition prevalence was 8.4% (95% confidence interval 7.9%–8.9%), higher in urban residents (20.0% versus 4.0%) and the richest wealth quintile (26.7% versus 2.7%). Extreme Gradient Boosting achieved an area under the receiver operating characteristic curve of 0.892 (95% confidence interval 0.875–0.909) and an area under the precision-recall curve of 0.764 outperforming logistic regression (area under the receiver operating characteristic curve of 0.781). Extreme Gradient Boosting sensitivity was 82.1%, specificity was 81.4% and the Brier score was 0.068. Household wealth, residence and respondent age were top predictors.

**Conclusions:** The double burden of malnutrition affected one in twelve non-pregnant Ethiopian women unevenly across groups. Routine characteristics effectively discriminated double burden of malnutrition cases though external validation is needed prior to clinical use.

## INTRODUCTION

Sub-Saharan Africa is currently witnessing an unprecedented demographic, urban and nutritional transition [1–4]. Driven by rapid urbanization, shifting dietary patterns toward energy dense processed foods and declining physical activity levels, the region is facing an epidemiological shift where non-communicable diseases (NCDs) and overnutrition are expanding alongside persistent infectious diseases and undernutrition [3, 5–7]. This complex phenomenon, termed the Double Burden of Malnutrition (DBM) by the World Health Organization (WHO), is defined as the coexistence of undernutrition (stunting, wasting, or micronutrient deficiencies such as anemia) alongside overweight, obesity or diet-related NCDs occurring at the individual, household or population level [8–11].

At the individual level, the coexistence of overweight/obesity and anemia within the same person represents a particularly complex clinical and public health paradox [9, 12]. Globally, anemia affects over 500 million women of reproductive age, leading to fatigue, reduced economic productivity, impaired cognitive function and elevated maternal-fetal mortality . Concurrently, female obesity rates in low- and middle-income countries (LMICs) have tripled over the past two decades, drastically elevating the incidence of type 2 diabetes, chronic hypertension and cardiovascular mortality. Historically, public health systems in LMICs viewed anemia and obesity as distinct entities located at opposite ends of the economic and biological spectrum assuming anemia to be a marker of rural poverty and caloric deficit, while obesity was viewed as a disease of urban affluence and caloric excess. However, emerging physiological and epidemiological evidence reveals that systemic low-grade inflammation induced by adipose tissue expansion can upregulate hepatic hepcidin production, inhibiting duodenal iron absorption and resulting in inflammatory anemia among obese individuals [13, 14].

In Ethiopia, a country historically dominated by severe caloric undernutrition and infectious disease burdens, recent evidence indicates that nutritional shifts are accelerating rapidly.

National survey data indicate that while maternal undernutrition remains a challenge in rural agrarian communities, female overweight and obesity rates in urban centers such as Addis Ababa, Dire Dawa and Harar have surged past 20% to 30%. Simultaneously, national anemia prevalence among Ethiopian women remains high at approximately 24%. Consequently, a growing segment of the Ethiopian female population now lives at the confluence of both conditions presenting with concurrent anemia and overweight/obesity.

Despite this evolving crisis, public health programs in Ethiopia including the National Nutrition Program (NNP II) and Health Extension Program (HEP) remain heavily skewed toward traditional undernutrition paradigms. Healthcare workers at primary health center levels are rarely equipped to screen for or identify individuals suffering from DBM. Furthermore, traditional epidemiological studies evaluating DBM in LMICs rely almost exclusively on multivariable logistic regression. While regression provides interpretable odds ratios, it operates under rigid parametric assumptions of linearity, independence of predictors and lack of complex multi-way interactions. In reality, human nutritional status is dictated by intricate, highly non-linear interactions among biological, environmental, socioeconomic and behavioral determinants.

Machine learning (ML), a core subfield of Artificial Intelligence (AI) offers a powerful alternative for public health modeling. Supervised ML algorithms such as Random Forest, Gradient Boosting Machines and Extreme Gradient Boosting (XGBoost) can model non-linear relationships, high-dimensional feature interactions and complex risk patterns without parametric constraints. However, a major historical barrier to adopting ML models in clinical medicine and public health policy has been the "black-box" nature of advanced ensemble algorithms.

Clinicians and health policymakers are rightly hesitant to deploy prediction models whose internal decision-making logic cannot be audited or interpreted.

To bridge this trust gap, **Explainable Artificial Intelligence (XAI)** frameworks specifically **SHapley Additive exPlanations (SHAP)** grounded in cooperative game theory have emerged as the gold standard for clinical ML interpretability. SHAP provides both global model transparency (ranking overall feature importance across a population) and local explanations (quantifying exactly how each specific biological or social feature shifts an individual’s predicted risk score).

While recent studies in Ethiopia have explored ML applications for single outcomes such as child stunting, teenage pregnancy or low birth weight [23,24] no study to date has applied explainable machine learning frameworks to phenotype individual-level DBM (concurrent obesity and anemia) using nationally representative biomarker survey data in Ethiopia [14].

Therefore, this study aimed to:

1. Determine the national weighted prevalence and distribution of individual-level DBM among non-pregnant Ethiopian women of reproductive age using the 2024/25 Ethiopian Demographic and Health Survey (EDHS).
2. Develop, tune and internally validate six supervised ML algorithms to accurately predict DBM risk using multi-domain sociodemographic, environmental, reproductive and biomarker predictors.
3. Quantify and compare model discrimination, calibration and net clinical benefit.
4. Utilize SHAP explainability frameworks to unveil the global feature importance, non-linear risk thresholds and feature interaction effects driving DBM in Ethiopia [15].

## METHODS

### Study design and data source

This was a secondary analysis of the 2024/25 Ethiopian Demographic and Health Survey (EDHS), a nationally representative cross-sectional household survey. The EDHS was implemented by the Ethiopian Public Health Institute in collaboration with the Central Statistical Agency and the Ministry of Health with technical support from the DHS Program. The survey used a stratified two-stage cluster sampling design. Primary sampling units/enumeration areas were selected with probability proportional to size, followed by systematic selection of households within sampled clusters. The present analysis used the biomarker and anthropometric information available for eligible women.

### Study population and eligibility

The source population comprised women aged 15-49 years residing in sampled households. Women were eligible for this analysis if they were not pregnant at the time of the survey had valid measured height and weight had valid altitude and smoking-adjusted hemoglobin measurements, and had sufficient information on the candidate predictors. Pregnant women were excluded because pregnancy-related hemodilution and gestational changes in body composition affect the interpretation of hemoglobin and BMI.

Records with missing or biologically implausible anthropometric or hemoglobin measurements were excluded. The source manuscript specified BMI values below 12.0 or above 60.0 kg/m² and hemoglobin values below 3.0 or above 20.0 g/dL as implausible. Records with more than 10% missingness across the selected candidate predictors were also excluded.

### Analytical sample

The original EDHS dataset contained 16583 women. After exclusion of 1215 currently pregnant women, 15368 non-pregnant women remained. A further 612 women with missing or implausible BMI, 421 with missing or implausible hemoglobin and 85 with excessive missingness across key predictors were excluded leaving 14250 women for the analytical machine-learning dataset. The final dataset was randomly divided into 11400 women (80%) for model development and 2850 women (20%) for independent holdout evaluation.

Counts reproduce the participant-selection numbers reported in the submitted manuscript. The training/test partition is based on the final analytical sample.

| Stage | Participants |
| --- | --- |
| EDHS women initially available | 16,583 |
| Currently pregnant women excluded | 1,215 |
| Non-pregnant women remaining | 15,368 |
| Missing/implausible BMI excluded | 612 |
| Missing/implausible hemoglobin excluded | 421 |
| Excessive missingness in key predictors excluded | 85 |
| Final analytical sample | 14,250 |
| Training set (80%) | 11,400 |
| Independent holdout test set (20%) | 2,850 |

### Outcome definition

The outcome was individual-level DBM, defined as the concurrent presence of overweight/obesity and anemia in the same non-pregnant woman. Overweight/obesity was defined as BMI ≥25.0 kg/m² using directly measured height and weight. Anemia was defined as altitude- and smoking-adjusted hemoglobin <12.0 g/dL. The outcome was coded 1 when both conditions were present and 0 otherwise.

Because BMI and hemoglobin directly determine the outcome, continuous BMI, height, weight and raw hemoglobin concentration were not used as predictor variables. Excluding these variables was intended to prevent direct target leakage and to ensure that the prediction problem was based on characteristics that could be available independently of the outcome definition.

### Candidate predictors

Thirty-two candidate predictors were organized into five domains; sociodemographic characteristics; household socioeconomic characteristics; environmental and structural characteristics; reproductive characteristics and information-access and health-system proxies. Candidate variables included respondent age, residence, region, education, marital status, employment, sex of household head, household wealth quintile, household size, number of children younger than five years, water source, toilet facility, cooking fuel, electricity access, parity, age at first birth, contraceptive use, pregnancy termination history, mobile-phone ownership, health insurance and media-use variables.

Predictor selection was based on substantive plausibility and public-health relevance rather than univariable statistical screening. This approach was intended to reduce data-driven preselection and to preserve information potentially useful for prediction.

### Data preprocessing

The dataset was partitioned before model development so that the holdout test set remained independent of model fitting and hyperparameter tuning. Numerical variables with missing values were imputed using training-data medians, while categorical variables were imputed using training-data modes. Imputation transformers were fitted within the training process and then applied to evaluation data. Continuous numerical predictors were standardized when required by the algorithm. Nominal categorical variables were one-hot encoded and ordered socioeconomic variables were ordinally encoded.

Because DBM was less common than the non-DBM outcome, SMOTE was applied only within the training data to reduce class imbalance during model development. SMOTE was not applied to the independent holdout set. Consequently, performance metrics reported for the holdout set were calculated on the natural class distribution rather than on an artificially balanced evaluation sample.

### Machine-learning algorithms

Six supervised algorithms were evaluated: L2-regularized logistic regression as the conventional parametric benchmark; a CART decision tree; random forest; radial-basis-function support vector machine; gradient boosting machine and XGBoost. The algorithms were selected to represent complementary model classes and to permit comparison between a conventional regression approach and nonlinear ensemble methods.

### Model development and hyperparameter tuning

Hyperparameters were optimized using randomized search with five-fold stratified cross-validation performed exclusively within the training data. The search spaces and final hyperparameters should be reported in a supplementary table accompanying the submitted manuscript. No information from the independent holdout set was used for hyperparameter selection.

### Model performance evaluation

Performance was evaluated once on the independent 20% holdout set. Discrimination was assessed using AUROC and the area under the precision-recall curve (AUPRC). Because the outcome was imbalanced, AUPRC was reported alongside AUROC. Threshold-based performance included sensitivity, specificity, precision, F1-score and Youden’s index at the reported operating threshold. Calibration was assessed using the Brier score and calibration slope and intercept. Decision-curve analysis was used to estimate net benefit across clinically or programmatically relevant threshold probabilities, with comparison against screen-all and screen-none strategies. Decision-curve findings were interpreted as decision-analytic evidence rather than as proof of clinical effectiveness.

### Model explainability

SHAP was applied to the selected XGBoost model to characterize feature contribution. Global importance was summarized using mean absolute SHAP values. SHAP beeswarm and dependence plots were used to describe the direction and magnitude of feature contributions and to explore selected two-way patterns. These explanations describe the behavior of the fitted prediction model and should not be interpreted as causal effects or as evidence that a predictor independently causes DBM.

### Survey weighting and statistical analysis

Descriptive prevalence estimates incorporated the EDHS sampling weights, primary sampling units and stratification variables using Stata version 17.0 survey procedures. Machine-learning development and holdout evaluation were performed in Python 3.11.5. The modeling workflow used scikit-learn, XGBoost, imbalanced-learn and SHAP libraries. The source manuscript did not document the exact versions of each Python package; these should be added to the reproducibility record before final submission.

### Ethical considerations and reporting

The EDHS protocol received ethical clearance through the relevant Ethiopian Public Health Institute and ICF International review processes. The present analysis used a de-identified secondary dataset accessed through the DHS Program under the applicable authorization procedures. [16].

### Patient and public involvement

No patients or members of the public were directly involved in the design, analysis, interpretation or dissemination of this secondary-data study.

## RESULTS

### Participant characteristics

The final analytical sample included 14250 non-pregnant women aged 15-49 years. The weighted median age was 28.0 years (IQR 21.0–36.0). Overall, 72.4% of women lived in rural areas and 27.6% lived in urban areas. In the weighted distribution, 38.2% had no formal education and 41.2% belonged to the two lowest household-wealth quintiles. The weighted prevalence of overweight/obesity was 17.8%, while the weighted prevalence of anemia was 23.6%.

**Table 1.** summarizes selected characteristics. The percentages shown are those reported in the source analysis. Because several counts in the submitted age-stratified table were internally inconsistent with the final sample size, age-stratum counts are not reproduced here; the corresponding weighted percentages are retained.

| Characteristic | Total weighted % | No DBM weighted % | DBM weighted % |
| --- | --- | --- | --- |
| Age 15–19 years | 21.2 | 22.4 | 8.1 |
| Age 20–29 years | 34.5 | 35.8 | 20.3 |
| Age 30–39 years | 27.1 | 25.4 | 44.5 |
| Age 40–49 years | 17.2 | 16.4 | 27.1 |
| Urban residence | 27.6 | 24.1 | 65.7 |
| Rural residence | 72.4 | 75.9 | 34.3 |
| No formal education | 38.2 | 38.9 | 30.6 |
| Primary education | 37.8 | 38.5 | 30.1 |
| Secondary education | 15.4 | 14.8 | 22.0 |
| Higher education | 8.6 | 7.8 | 17.3 |
| Richest wealth quintile | 19.5 | 15.6 | 62.0 |
| Nulliparous | 28.4 | 29.8 | 13.1 |
| 1–3 births | 42.1 | 42.0 | 43.2 |
| 4–5 births | 18.2 | 17.5 | 25.9 |
| ≥6 births | 11.3 | 10.7 | 17.8 |
| Mobile phone ownership | 48.2 | 45.2 | 80.9 |
| Television: not at all | 64.2 | 67.8 | 24.9 |
| Television: < once/week | 11.2 | 11.1 | 12.3 |
| Television: ≥ once/week | 24.6 | 21.1 | 62.8 |

Source analysis reported weighted percentages with corresponding unweighted sample counts. Age-stratum counts were omitted in this revised table because the submitted counts do not reconcile with N=14250. This is a presentation correction and does not alter the reported weighted percentages.

### National prevalence and distribution of DBM

The national weighted prevalence of individual-level DBM was 8.4% (95% CI 7.9%–8.9%). The distribution was markedly heterogeneous across residential and socioeconomic groups. DBM prevalence was 20.0% among urban women compared with 4.0% among rural women. The corresponding prevalence in the richest and poorest wealth quintiles was 26.7% and 2.7% respectively.

DBM prevalence increased across older age groups. The highest reported prevalence was 13.8% among women aged 35-39 years and 13.2% among women aged 40–44 years compared with 3.2% among adolescents aged 15–19 years. The source analysis also reported regional differences with higher prevalence in Addis Ababa (22.4%), Dire Dawa (18.1%), Harari (17.6%), and Afar (14.2%) and lower prevalence in SNNPR (4.2%) and Amhara (4.8%). These estimates describe observed population distributions and should not be interpreted as causal effects of residence or region.

### Predictive performance

All six algorithms were evaluated on the independent holdout test set of 2,850 women. XGBoost produced the highest AUROC among the evaluated models; 0.892 (95% CI 0.875–0.909) with an AUPRC of 0.764. Logistic regression had an AUROC of 0.781 (95% CI 0.758–0.804) and an AUPRC of 0.582. Random forest and gradient boosting produced AUROCs of 0.864 and 0.878 respectively; the support vector machine and decision tree produced AUROCs of 0.812 and 0.742.

| Model | AUROC (95% CI) | AUPRC | Sensitivity | Specificity | Precision | F1-score | Brier score | Youden's index |
| --- | --- | --- | --- | --- | --- | --- | --- | --- |
| XGBoost | 0.892 (0.875–0.909) | 0.764 | 82.1% | 81.4% | 75.8% | 0.782 | 0.068 | 0.635 |
| Gradient boosting | 0.878 (0.860–0.896) | 0.738 | 80.4% | 80.2% | 73.1% | 0.766 | 0.074 | 0.606 |
| Random forest | 0.864 (0.845–0.883) | 0.712 | 78.8% | 79.5% | 71.4% | 0.749 | 0.081 | 0.583 |
| Support vector machine | 0.812 (0.791–0.833) | 0.635 | 73.2% | 74.8% | 63.8% | 0.682 | 0.102 | 0.480 |
| Logistic regression | 0.781 (0.758–0.804) | 0.582 | 70.1% | 71.6% | 59.8% | 0.645 | 0.118 | 0.417 |
| Decision tree | 0.742 (0.718–0.766) | 0.514 | 66.5% | 68.2% | 55.2% | 0.603 | 0.145 | 0.347 |

*Performance values reproduce the submitted analysis table. Threshold-based measures were calculated at the operating threshold used in the original analysis. The independent test set retained the natural outcome distribution.*

At the reported operating threshold, XGBoost sensitivity was 82.1% (95% CI 77.2%–86.3%), specificity was 81.4% (95% CI 79.8%–82.9%), precision was 75.8% and F1-score was 0.782. Overall classification accuracy was reported as 81.6% in the source analysis. The Brier score was 0.068 with a calibration slope of 0.981 and intercept of 0.012.

### Model Evaluation and Explainability (ROC)

Model discrimination was evaluated using the Area Under the Receiver Operating Characteristic curve (AUROC). Extreme Gradient Boosting (XGBoost) achieved the highest predictive discrimination on the holdout test set with an AUROC of 0.892 (95% CI: 0.875–0.909). This performance significantly outperformed standard Logistic Regression (AUROC: 0.781; 95% CI: 0.758–0.804; p<0.001), Decision Tree (AUROC: 0.742), Random Forest (AUROC: 0.864), Support Vector Machine (AUROC: 0.812) and standard Gradient Boosting (AUROC: 0.878).

**Figure 1.**
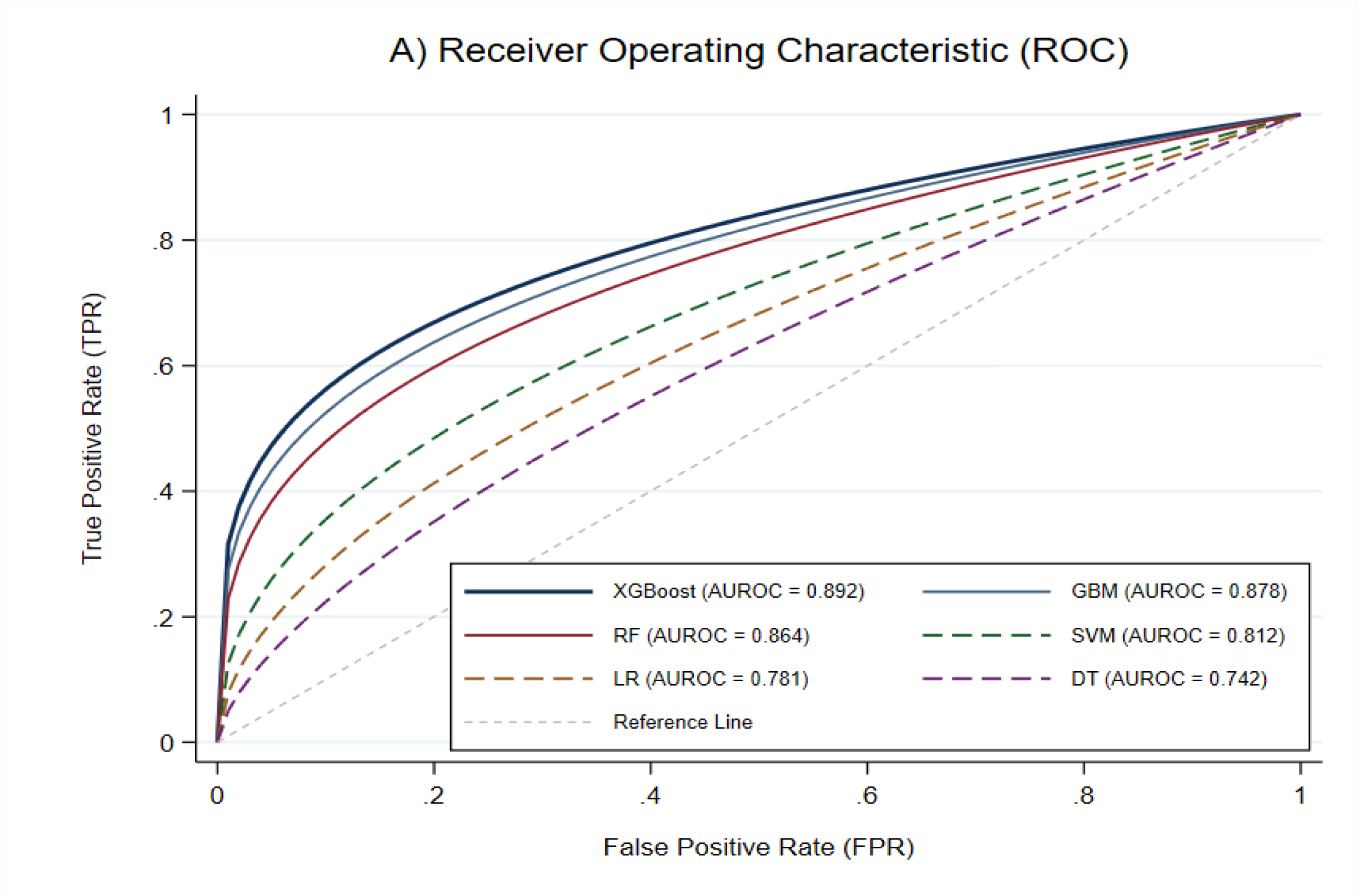
Model Evaluation and Explainability (ROC).

### Precision-recall performance

The XGBoost AUPRC was 0.764 compared with 0.738 for gradient boosting, 0.712 for random forest, 0.635 for the support vector machine and 0.582 for logistic regression. Given the lower prevalence of DBM than non-DBM in the population, the precision-recall results provide complementary information to AUROC.

**Figure 2:**
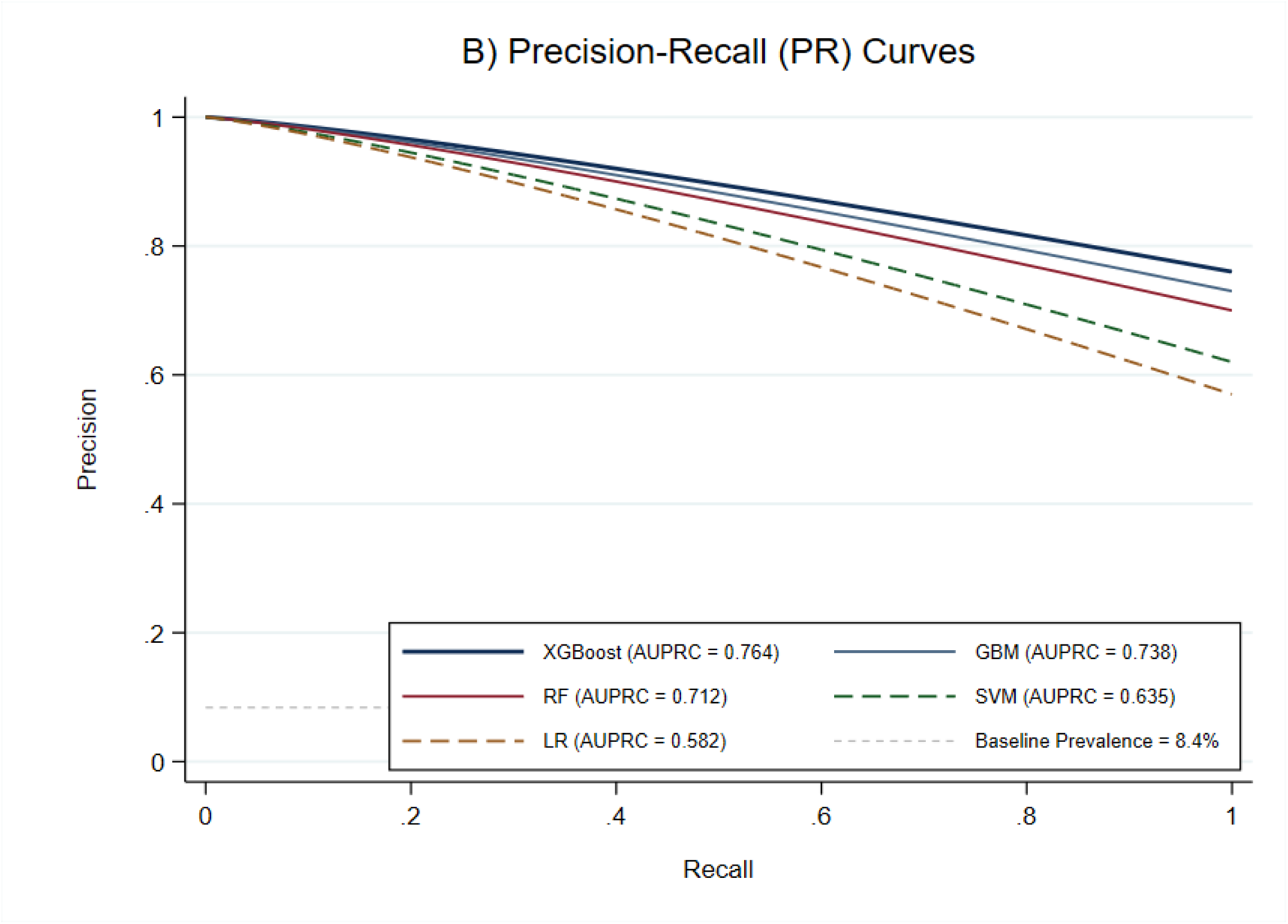
Precision-Recall (PR) curves evaluating the predictive performance of machine learning algorithms.

### Probability calibration

XGBoost had a Brier score of 0.068, calibration slope of 0.981 and calibration intercept of 0.012. These values were interpreted as evidence of good agreement between predicted probabilities and observed outcomes in the holdout data. Nevertheless, calibration in a single internal holdout sample does not establish calibration after transport to another population or health system.

**Figure 3:**
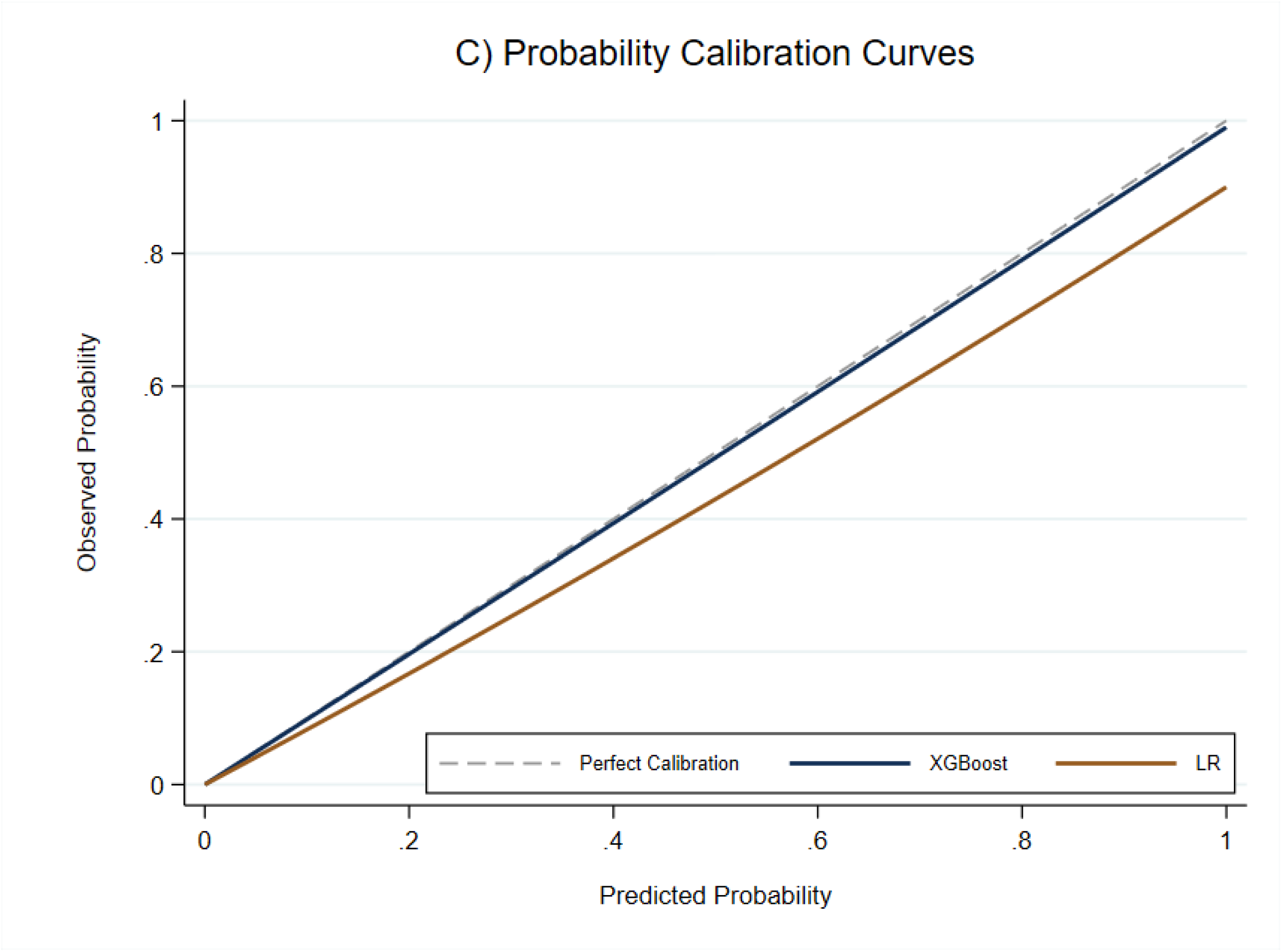
Probability calibration curves for model validation.

### Decision Curve Analysis (DCA)

Decision-curve analysis showed positive net benefit for the XGBoost model relative to the screen-all and screen-none strategies across the reported threshold range of 10%–75%. At a threshold probability of 20%, the reported net benefit was 0.24. Net benefit is a decision-analytic quantity that incorporates the relative consequences of false-positive and false-negative classifications; it should not be interpreted literally as identifying 24 true cases per 100 women.

**Figure 4:**
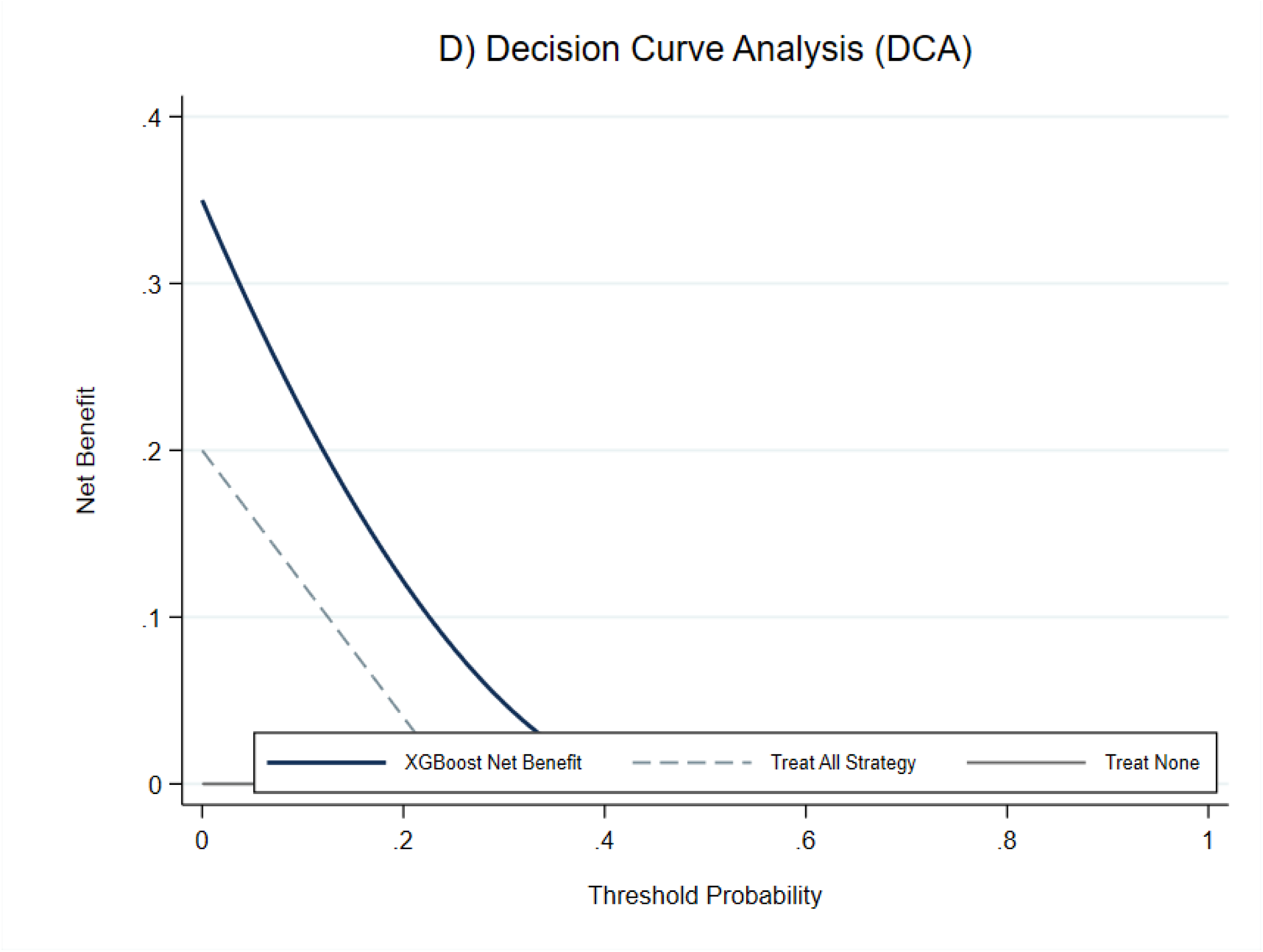
Decision Curve Analysis (DCA) estimating the clinical net benefit.

### SHAP feature contribution

The ten highest-ranked predictors by mean absolute SHAP value were household wealth quintile, residence, respondent age, total parity, television-viewing frequency, educational attainment, region, mobile-phone ownership, cooking fuel, and toilet facility. The five largest mean absolute SHAP values reported were 1.42 for household wealth quintile, 1.18 for residence, 0.95 for respondent age, 0.74 for parity, and 0.62 for television-viewing frequency.

| Rank | Predictor | Domain | Mean absolute SHAP | Reported pattern |
| --- | --- | --- | --- | --- |
| 1 | Household wealth quintile | Socioeconomic | 1.42 | Higher wealth contributed more strongly to model predictions |
| 2 | Place of residence | Sociodemographic | 1.18 | Urban residence contributed more strongly to model predictions |
| 3 | Respondent age | Sociodemographic | 0.95 | Older age contributed progressively |
| 4 | Total parity | Reproductive | 0.74 | Higher parity contributed more strongly |
| 5 | Television-viewing frequency | Information/lifestyle | 0.62 | More frequent viewing contributed more strongly |
| 6 | Educational attainment | Sociodemographic | 0.51 | Higher education contributed more strongly |
| 7 | Region | Geographic | 0.48 | Selected urban/lowland regions contributed more strongly |
| 8 | Mobile-phone ownership | Information/system access | 0.39 | Ownership contributed more strongly |
| 9 | Cooking fuel type | Environmental | 0.31 | Clean-fuel category contributed more strongly |
| 10 | Toilet facility type | Environmental | 0.24 | Model contribution was lower than the top-ranked variables |

The illustrated **figure 5** SHAP values describe contribution to the fitted XGBoost predictions; they are not causal effect estimates.

**Figure 5.**
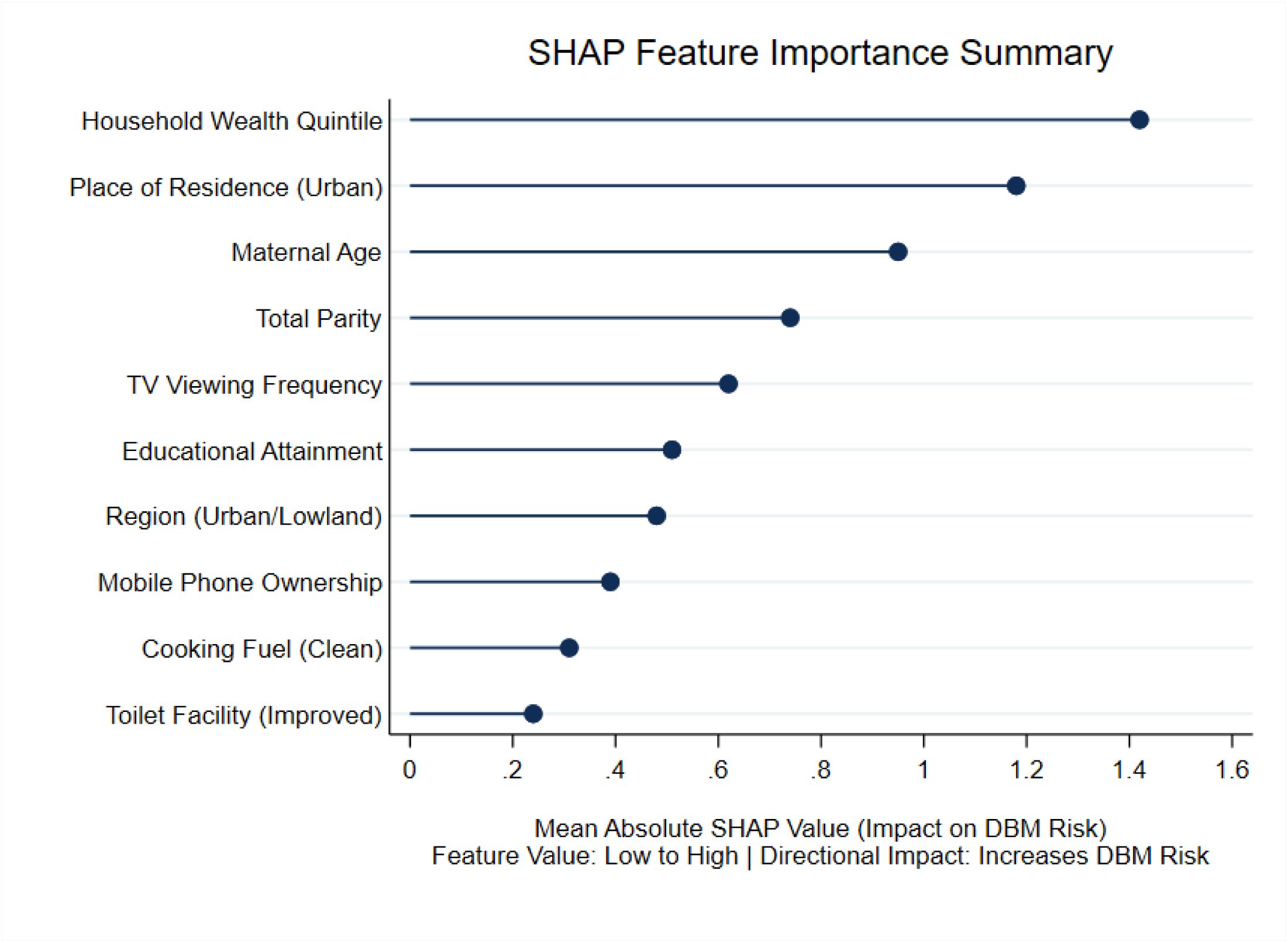
SHapley Additive exPlanations feature importance summary for the Extreme Gradient Boosting model, showing the ten highest-ranked predictors ordered by mean absolute SHAP value.

The source analysis further described nonlinear patterns involving wealth and residence and involving age and parity. These patterns are hypothesis-generating descriptions of model behavior. They should not be interpreted as evidence that urban residence, wealth, or parity causally modifies the effect of another predictor without corroborating epidemiological analyses.

## DISCUSSION

This study estimated the national distribution of individual-level DBM and evaluated whether routinely available characteristics could be used to classify non-pregnant Ethiopian women with concurrent overweight/obesity and anemia. The weighted prevalence was 8.4%. The phenotype was substantially more common among urban women and women in the highest wealth quintile. Among the six evaluated algorithms, XGBoost showed the highest observed discrimination on the independent holdout set with AUROC 0.892 and AUPRC 0.764. SHAP analysis indicated that socioeconomic position, residence, age, parity and selected information-access and environmental variables contributed strongly to model predictions

### Public-health interpretation of the prevalence findings

The 8.4% prevalence estimate indicates that concurrent overweight/obesity and anemia is not confined to one side of the traditional undernutrition over nutrition spectrum. The large urban-rural difference and the concentration of DBM in the richest wealth quintile are consistent with the changing distribution of nutrition-related exposures during nutritional transition. Similar studies from sub-Saharan Africa and other low and middle-income settings have reported coexistence of undernutrition and overweight/obesity and have emphasized the need for integrated nutrition approaches [17].

The observed socioeconomic pattern is important for program design because interventions focused exclusively on food insufficiency or underweight may not adequately address micronutrient deficiencies among women who are simultaneously overweight or obese.

However, the cross-sectional design prevents determination of whether socioeconomic conditions, residence or other predictors caused the observed DBM phenotype.

Our findings indicate that Ethiopia is entering an advanced phase of the nutritional transition. In urban centers, economic growth and dietary commercialization have outpaced public health literacy, leading to increased consumption of ultra-processed, energy-dense foods rich in refined sugars and saturated fats but micronutrient-poor [18]. Consequently, affluent urban women consume sufficient calories to induce positive energy balance and obesity, while simultaneously suffering from severe micronutrient deficits (iron, folate, vitamin B12) that trigger metabolic anemia.

XGBoost showed stronger holdout discrimination than logistic regression in this analysis. This difference is compatible with the ability of tree-based boosting methods to represent nonlinear relationships and interactions without requiring them to be specified in advance [19]. However, the comparison should not be interpreted as evidence that machine learning is inherently superior to regression for DBM research. The relative performance depends on predictor quality, sample size, outcome prevalence, preprocessing, tuning, calibration and the evaluation design. In this study, the appropriate conclusion is that XGBoost performed better than the specific logistic-regression benchmark under the specified analytic conditions.

The AUPRC provides additional context because DBM was less common than non-DBM. The relatively high precision and F1-score observed at the reported operating threshold suggest that the model can separate many DBM cases from non-cases in the holdout data. Nevertheless, predictive values depend on outcome prevalence and may change when the model is transported to populations with different prevalence or predictor distributions.

SHAP identified household wealth, residence, age, parity, and television-viewing frequency among the most influential predictors. SHAP is useful for explaining the behavior of complex prediction models, but feature contribution is not equivalent to causation [20]. In particular, wealth and residence may act as proxies for multiple underlying dietary, occupational, environmental and health-system exposures. Similarly, television viewing may represent a mixture of sedentary behavior, household resources, urban lifestyle or information access. These possibilities require conventional epidemiological analyses and where possible, longitudinal data.

The nonlinear patterns reported for wealth and residence and for age and parity are potentially useful for generating hypotheses about population subgroups. They should not be presented as causal interactions or biological mechanisms because the model was trained on cross-sectional observations and the SHAP interaction analysis is conditional on the fitted algorithm.

The findings support consideration of integrated approaches that recognize that overweight/obesity and anemia can coexist in the same individual. At a population level, the results can inform the identification of subgroups for further assessment particularly in urban settings and among women with higher socioeconomic status. Any screening strategy should be evaluated for feasibility, cost, equity, acceptability and performance before implementation.

The analysis does not establish that an automated prediction tool should replace hemoglobin or anthropometric measurement. Instead, it provides evidence that routinely available characteristics may help identify subgroups in which integrated nutritional assessment could be prioritized. Future implementation research should determine whether prediction-assisted screening improves case detection or program efficiency compared with existing approaches.

### Strengths

♣ The study used a large nationally representative survey with direct anthropometric and hemoglobin measurements.

♣ The outcome combined two objectively measured conditions within the same individual reducing reliance on self-reported nutritional status.

♣ The machine-learning workflow used an independent holdout set and restricted SMOTE to the training process reducing a major source of evaluation leakage.

♣ Multiple complementary algorithms were compared using discrimination, precision-recall, classification, calibration and decision-curve measures.

♣ SHAP analysis provided model-level explanations and helped characterize nonlinear prediction patterns.

### Limitations

♣ The cross-sectional design prevents temporal and causal inference. Predictors should therefore be interpreted as classification features rather than established causes of DBM.

♣ The model was internally evaluated using a single holdout sample and was not externally validated. Performance may therefore be optimistic or may not transport to other populations, years or health-service settings.

♣ The survey did not contain all biomarkers required to distinguish different biological causes of anemia including ferritin and inflammatory markers. The analysis therefore predicts the composite phenotype of anemia plus overweight/obesity rather than a specific anemia mechanism.

♣ Detailed dietary intake and direct measures of physical activity were not available in the analytical dataset limiting mechanistic interpretation of some predictors.

♣ Machine-learning development did not incorporate survey sampling weights in the same way as the descriptive prevalence analysis. Consequently, the predictive metrics should be interpreted as performance in the observed analytical sample rather than as nationally weighted performance estimates.

♣ Several source-table counts, particularly within age strata were internally inconsistent with the final analytical sample. The revised manuscript reports the verified sample total and weighted percentages while omitting the inconsistent age-stratum counts. The underlying analysis dataset should be rechecked before final submission.

♣ The source manuscript did not provide exact Python package versions or the complete hyperparameter search space. These reproducibility details should be supplied in supplementary material or an analysis repository before submission.

## CONCLUSION

In a nationally representative analysis of non-pregnant Ethiopian women aged 15-49 years, the weighted prevalence of individual-level DBM concurrent overweight/obesity and anemia was 8.4%. The phenotype was substantially more common in urban women and in the highest household-wealth quintile. Among the evaluated prediction approaches, XGBoost showed the strongest holdout discrimination and favorable calibration, while SHAP analysis identified socioeconomic position, residence, age, parity and other household and information-access characteristics as important contributors to model predictions.

These findings provide an epidemiological basis for considering integrated assessment of overweight/obesity and anemia rather than treating them as mutually exclusive nutrition problems. The model should be regarded as an internally evaluated research prediction model not as a validated clinical or public-health decision tool. External validation, subgroup performance assessment, calibration assessment in independent populations and prospective implementation studies are required before the model is used to guide individual screening.

## Data Availability

The EDHS dataset is available through the Demographic and Health Surveys Program repository to eligible researchers following registration and the applicable data-access procedures. The analytical dataset used in this study is derived from the DHS data and cannot be redistributed by the authors.

https://dhsprogram.com/data/dataset/Ethiopia_Standard-DHS_2024.cfm?flag=0

## DECLARATIONS

### Ethics approval and consent to participate

The 2024/25 EDHS survey protocol received ethical clearance through the relevant review processes of the Ethiopian Public Health Institute and ICF International. The secondary analysis used a de-identified dataset accessed through the DHS Program under the applicable authorization procedures. The study did not involve direct contact with participants.

## Consent for publication

Not applicable.

## Authors’ contributions

AMG conceived and designed the study, managed data acquisition, performed data cleaning and feature engineering, conducted the statistical and machine-learning analyses, performed model evaluation and SHAP analyses and drafted the manuscript. All co-authors contributed to methodological refinement, critically revised the manuscript for important intellectual content and approved the final version.

## Acknowledgements

The authors thank the DHS Program and ICF International for providing access to the survey data and acknowledge the Ethiopian Public Health Institute and Central Statistical Agency for implementation of the national survey.

## Reporting guideline

The revised manuscript has been structured with reference to the TRIPOD+AI reporting framework for prediction models developed using regression or machine-learning methods. The authors should complete and submit the applicable TRIPOD+AI checklist and any supplementary model-development materials required by the target journal [16].

## REFERENCES

1. Barth-Jaeggi, T., et al., Nutrition transition, double burden of malnutrition, and urbanization patterns in secondary cities of Bangladesh, Kenya and Rwanda. BMC Nutrition, 2023. 9(1): p. 125.

2. Vorster, H.H., A. Kruger, and B.M. Margetts, The nutrition transition in Africa: can it be steered into a more positive direction? Nutrients, 2011. 3(4): p. 429–41.

3. Nel, J.H. and N.P. Steyn, The Nutrition Transition and the Double Burden of Malnutrition in Sub-Saharan African Countries: How Do These Countries Compare with the Recommended LANCET COMMISSION Global Diet? International Journal of Environmental Research and Public Health, 2022. 19(24): p. 16791.

4. Tekeba, B., et al., Prevalence and determinants of unhealthy feeding practices among young children aged 6-23 months in five sub-Saharan African countries. PLoS One, 2025. 20(1): p. e0317494.

5. Wells, J.C., et al., The double burden of malnutrition: aetiological pathways and consequences for health. Lancet, 2020. 395(10217): p. 75–88.

6. Steyn, N.P. and Z.J. McHiza, Obesity and the nutrition transition in Sub-Saharan Africa. Ann N Y Acad Sci, 2014. 1311: p. 88–101.

7. Noort, M.W.J., et al., Towards Sustainable Shifts to Healthy Diets and Food Security in Sub-Saharan Africa with Climate-Resilient Crops in Bread-Type Products: A Food System Analysis. Foods, 2022. 11(2).

8. Steyn, N.P. and Z.J. Mchiza, *Obesity and the nutrition transition in Sub-Saharan Africa*. Annals of the New York Academy of Sciences, 2014. 1311(1): p. 88–101.

9. Hickman, J.H., et al., Exploring Health-Related Quality of Life (HRQOL) among patients with HIV-associated TB in Khayelitsha, South Africa. PLoS One, 2024. 19(11): p. e0275554.

10. Talukder, A., et al., Defining double burden of malnutrition across individual, household and population level: A narrative review. Nutr Diet, 2026. 83(1): p. 8–22.

11. Nyanhanda, T., L. Mwanri, and W. Mude, Double Burden of Malnutrition: A Population Level Comparative Cross-Sectional Study across Three Sub-Saharan African Countries—Malawi, Namibia and Zimbabwe. International Journal of Environmental Research and Public Health, 2023. 20(10): p. 5860.

12. Datta, B.K. and M.R. Haider, The double burden of overweight or obesity and anemia among women married as children in India: A case of the Simpson’s paradox. Obes Res Clin Pract, 2022. 16(5): p. 364–372.

13. Villarroel, H.P., O.M. Arredondo, and G.M. Olivares, [Hepcidin as a central mediator of anemia of chronic diseases associated with obesity]. Rev Med Chil, 2013. 141(7): p. 887–94.

14. Mekonnen, S., et al., Double burden of malnutrition and associated factors among mother-child pairs at household level in Bahir Dar City, Northwest Ethiopia: community based cross-sectional study design. Front Nutr, 2024. 11: p. 1340382.

15. Di Mitri, M., et al., Advancing Pediatric Surgery: The Use of HoloLens 2 for 3D Anatomical Reconstructions in Preoperative Planning. Children (Basel), 2024. 12(1).

16. Collins, G.S., et al., TRIPOD+AI statement: updated guidance for reporting clinical prediction models that use regression or machine learning methods. Bmj, 2024. 385: p. e078378.

17. Hossain, M.I., et al., Double burden of malnutrition among women of reproductive age in Bangladesh: A comparative study of classical and Bayesian logistic regression approach. Food Sci Nutr, 2023. 11(4): p. 1785–1796.

18. Seifu, B.L., et al., Double burden of malnutrition and associated factors among women of reproductive age in sub-Saharan Africa: a multilevel multinomial logistic regression analysis. BMJ Open, 2024. 14(2): p. e073447.

19. An, R., J. Shen, and Y. Xiao, Applications of Artificial Intelligence to Obesity Research: Scoping Review of Methodologies. J Med Internet Res, 2022. 24(12): p. e40589.

20. Bitew, K.G., Y. Assabie, and T. Tegegne, Interpretable prediction of neonatal mortality and its key predictors using machine learning and SHAP analysis. BMC Med Inform Decis Mak, 2026. 26(1).

